# Common and separable neurofunctional dysregulations characterize obsessive-compulsive, substance use, and gaming disorders – evidence from an activation likelihood meta-analysis of functional imaging studies

**DOI:** 10.1101/2020.05.21.20108316

**Authors:** Benjamin Klugah-Brown, Xinqi Zhou, Basant K. Pradhan, Jana Zweerings, Klaus Mathiak, Bharat Biswal, Benjamin Becker

## Abstract

**Background:** Compulsivity and loss of behavioral control represent core symptoms in obsessive-compulsive disorder (OCD), substance use disorder (SUD), and internet gaming disorder (IGD). Despite animal models suggesting compulsivity mediated by cortico-striatal circuits and several neuroimaging case-control studies positing common/distinct neurofunctional alterations in these disorders a systematic examination is still lacking.

**Aims:** The present study capitalized on previous case-control fMRI studies to determine shared and disorder-specific neurofunctional alterations among three disorders.

**Methods:** Task-based fMRI studies in SUD, OCD, and IGD were obtained. Coordinate-based meta-analyses were performed within each disorder. Next contrast and conjunction meta-analyses were done to determine differential and common neurofunctional alterations between the disorders. Task-paradigm were group according to RDoC domains to determine contributions of underlying behavioral domains. Find pre-registration of the study here (https://osf.io/j8wct/).

**Results:** 144 articles were included representing 6897 individuals (SUD=2418, controls=2332; IGD=361, controls=360; OCD=715, controls=711). Conjunction meta-analyses revealed shared alterations in anterior insular cortex between OCD, and pooled as-well-as separate SUDs. SUD exhibited pronounced dorso-striatal alterations as compared to both, OCD and IGD. IGD shared frontal, particularly cingulate alterations with all SUDs. IGD demonstrated temporal alterations compared to both, SUD and OCD. No robust overlap between IGD and OCD was observed. Across the disorders, neurofunctional alterations were mainly contributed to by cognitive systems and positive valence RDoC domains.

**Conclusion:** The present findings indicate that neurofunctional dysregulations in prefrontal regions engaged in regulatory control share neurofunctional alterations across substance and behavioral addictions, while shared neurofunctional dysregulations in the anterior insula may mediate compulsivity in substance addiction and obsessive-compulsive disorders.

## Introduction

Compulsivity refers to maladaptive repetitive and irrational behaviors, which are driven by strong urges in the context of a loss of behavioral control and which are repeated despite adverse consequences. Compulsivity is a core symptom of obsessive-compulsive disorder (OCD) characterized by intrusive thoughts and an uncontrollable urge to perform repetitive rituals but is also increasingly recognized as a core symptomatic domain in substance and behavioral addictions (1). In addictive disorders, the transition from initial voluntary and impulsive use towards compulsive use has been considered as a core pathological mechanism promoting the loss of behavioral control and continued use despite negative consequences (2). In line with the symptomatologic overlap and the course of the development of the Research Domain Criteria (RDoC) framework overarching conceptualizations have proposed that compulsivity may represent a transdiagnostic dimension across these disorders (1,3–5).

Animal models and experimental research in humans suggest that dysregulations in habit formation and reward processing, as well as impaired regulatory control and cognitive flexibility, promote the development of compulsive behavior (5,6). In line with these conceptualizations previous meta-analyses of case-control studies comparing either patients with OCD or substance use disorders (SUD) with healthy controls reported partly overlapping dysregulations in the domains of reward-related processes, associative learning and goal-directed behavior underlying habit formation as well as marked deficits in executive control and cognitive flexibility (7–12). On the neural level compulsivity has been tightly linked to the structural and functional integrity of segregated yet interacting cortico-striatal loops that neurally mediate habit formation, reward processing and executive control (13,14). In line with elaborated animal models of compulsive and addictive behavior (15,16) a growing number of recent meta-analysis covering case-control studies in either OCD and SUD have demonstrated robust and partly overlapping neurofunctional alterations in the intrinsic communication and task-related activation in cortico-striatal circuits (17–21), suggesting partly overlapping neuropathological pathways between these disorders.

Based on increasing prevalence rates (22,23), internet gaming disorder (IGD) has been recently included as an emerging behavioral addiction in the Diagnostic and Statistical Manual of Mental Disorders-Fifth Edition (DSM-5, APA) (24) and the International Classification of Diseases 11 (ICD 11, WHO) (25). The proposed IGD symptoms in both diagnostic systems are strongly aligned with symptoms for SUD and include compulsive and recurrent engagement in gaming and progressively impaired loss of behavioral control leading to continued gaming despite negative consequences (26). Based on the symptomatic overlap with SUDs, recent overarching models of IGD propose that the development of compulsive use and the loss of behavioral control in IGD are mediated by mal-adaptations in behavioral domains and cortico-striatal circuits that also drive the development of substance-based addictions (27,28). A rapidly growing number of experimental studies suggest functional impairments in IGD, including behavioral characteristics of compulsivity (29–31), deficient executive and inhibitory control (32) and to a lesser extend alterations in reward processing (33). Previous meta-analyses covering case-control functional imaging studies in subjects with problematic internet use and IGD suggest robust neurofunctional alterations during cognitive and reward processing in frontal and insular regions, yet less robust evidence for functional alterations in striatal regions (34–36).

Despite overlapping compulsive symptoms between IGD, SUD and OCD on the clinical level and initial studies suggesting that compulsivity across these disorders is driven by shared dysregulations in underlying behavioral domains (31), common and distinct neurobiological mal-adaptations between these disorders have not been systematically determined. While compulsivity is not an explicit dimensional construct or domain in the RDoC framework, it can be conceptualized by various RDoC domains, specifically dysregulations in cognitive systems as well as domains included in the positive valence domain. Moreover, the RDoC framework implements a dimensional contract approach that allowed us to account for the specific task-paradigms used in the original studies and thus allowed us to capitalize on sufficient power to determine shared and distinct neurofunctional alterations (for a similar integration of the RDoC approach with task-fMRI meta-analysis see (37)).

Summarizing, compulsivity is an increasingly recognized transdiagnostic domain underlying behavioral dysregulation across IGD, SUD, and OCD. Against this background the present comparative meta-analysis aimed at determining common neural alterations in these disorders by capitalizing on previous fMRI case-control studies conducted in individuals with obsessive-compulsive disorder (OCD), substance use disorders (SUD) and internet gaming disorder (IGD). Based on the currently prevailing disorder models and the previous literature we hypothesize shared transdiagnostic alterations in frontal and striatal regions.

## Methods

This meta-analysis adheres to Preferred Reporting Items for Systematic Reviews and Meta-Analyses (PRISMA) ((38)) guidelines and the principles of conducting coordinate-based meta-analyses ((39)). The meta-analytic approach and analyses were pre-registered on the Open Science Framework (DOI 10.17605/OSF.IO/J8WCT, https://osf.io/j8wct/). We initially conducted a literature search on task-based fMRI studies in SUD, IGD, and OCD using four relevant databases and additionally identified relevant studies in pertinent reviews. Titles and abstracts returned by the search were examined for subsequent full-text screening and inclusion. The screening process resulted in a total of 152 peer-reviewed original articles of which 99, 35 and 18 employed case-control designs in SUD, OCD, and IGD respectively. Articles that passed the inclusion criteria were further grouped based on the task paradigm according to RDoC domains as described e.g. in ((3)). Finally, coordinates from the original studies in the three diagnostic categories were extracted and analyzed utilizing activation likelihood estimation (ALE) (40).

### Literature search and databases

Task-based fMRI studies reporting results on the whole-brain level, employing case-control designs in SUD, OCD and IGD samples that were published in English language between January 1^st^, 2000 to January 31^st^, 2020 were included. The following search terms were applied: “functional magnetic resonance imaging” OR “fMRI” AND “substance use disorder” OR “obsessive-compulsive disorder” OR “internet gaming disorder” on four databases (PubMed, Web of Science, Neurosynth and Scopus). Only articles with case-control designs reporting differences between the respective diagnostic group with a healthy reference group were included. Additional exclusion criteria were as follows: 1. Articles reporting only region-of-interest (ROI) results (if the study additionally reported whole-brain corrected findings these were included), 2. Articles with poly-drug users or conducted in samples with comorbidity (severe psychiatric or somatic disorders, e.g. schizophrenia or HIV), 3. Articles focusing on parental substance use exposure, and 4. Articles reporting results from the same dataset previously published. A flow chart depicting the article selection for the main and sub-meta-analysis is shown in **Fig 1**.

**Figure 1.**
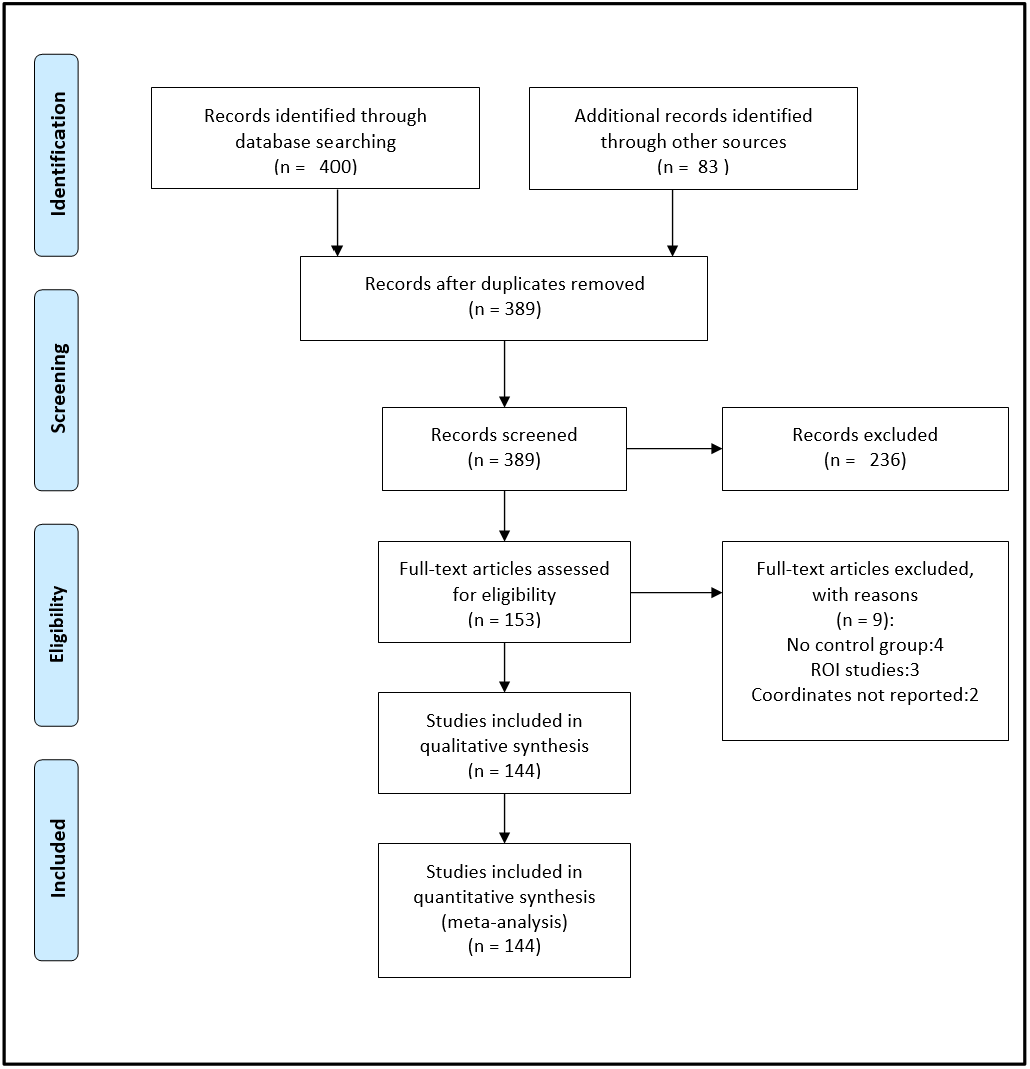
Flow-chart of the selection procedure. Number of experiments included in the analysis of interest.

### Activation likelihood estimation (ALE)

All coordinates (either provided in Talairach or Montreal Neurological Institute, MNI, standard space) and sample size were manually extracted from the original articles and next subjected to the current version of ALE (40–42) for meta-analytic analyses. ALE uses the convergence of whole-brain coordinates from experiments under a non-random spatial distribution to create clusters formed by equal probabilities of voxels. The ALE map indicates clusters of convergence. In line with recommendations, cluster-level familywise error-corrected (FWE) with a cluster forming threshold of p<0.05 and p<0.001 (43) respectively, were employed. The p-value accounts for the proportion of the random spatial relation between the various experiments under the null distribution. In line with the aim of the study, we mainly focused differences (contrasts) and overlap (conjunction) among the three diagnostic groups in the following way: first, ALEs were calculated separately for each diagnostic category and next the pooled coordinates between any two groups were created yielding six permutations. Finally, for the six permutations, three results are expected, i.e. one between each pair. At each computation, significant clusters were estimated by applying 5,000 Monte-Carlo simulations (44). Corresponding voxel probabilities, as well as the contribution from the RDoC domains, were extracted from the obtained modeled activation maps which were used for further analysis using the Kruskal-Wallis test.

### Contrast and conjunction analyses

The major aim of the present study was to determine divergent (disorder-specific) and convergent (transdiagnostic) neurofunctional alterations between the diagnostic categories in comparison to their respective healthy reference groups. Firstly, in the contrast analysis, we used the “contrast analysis” algorithm implemented in the ALE toolbox method to uncover significant clusters characterizing each diagnostic group. Next, using the corrected maps of the individual ALEs, voxel differences were computed to determine significant neurofunctional alterations within each disorder relative to the respective healthy reference group by means disorder-specific meta-analyses. Null distributions were generated from the ALE scores using random permutation of all experiments, after which the ALE score of the equal-sized group similar to the individual computed maps were randomly permuted for all voxels in the represented brain. Secondly, we aimed at determining divergent neurofunctional alterations between the disorder categories and thus computed differential contrasts between two of the diagnostic categories. Thirdly, we aimed to identify common neurofunctional alterations between diagnostic categories. To this end, we computed conjunctions between each of the two diagnostic categories. We computed the conjunction by taking the voxel-wise minimum value of the input ALE images. The expected results represent the intersections for shared neurofunctional alterations in these disorders. For all analyses a cluster-level FWE, with p< 0.05 and an initial cluster forming threshold p<0.001 was employed.

### Exploratory analysis

Firstly, based on our previous meta-analysis demonstrating robust and shared neurofunctional alterations in a core striato-frontal network across SUDs (Klugah-Brown et al., 2020) our primary analysis compared the pooled data from all SUDs with OCD and IGD respectively. However, to further account for SUD-specific alterations and the considerably higher number of SUD studies compared to OCD and IGD respectively we additionally computed separate conjunction analyses between each SUD (Cocaine, Cannabis, Alcohol and Nicotine) and IGD/OCD. Secondly, to further map the determined disorder-specific and common transdiagnostic alterations on the underlying behavioral domains the experimental paradigms were mapped onto the proposed RDoC domains, specifically the cognitive systems (n=63), positive valence systems (n=41), negative valence systems (n=14), cross-domain systems (14), and social processing (n=12) domains (for a detailed overview of specific task paradigms associated with RDoC domains, see the supplementary section of (3). Additionally, to test for effect sizes in publications, we computed Egger’s test within each of the diagnostic groups to determine heterogeneity (45). For each of the main, differential and conjunction effects, the probability of functional changes was extracted per-voxel and the contributions from each RDoC domain were subjected to Kruskal-Wallis tests (Bonferroni corrected for the number of comparisons). This analysis aimed at resolving a potential bias related to different task paradigms employed in the diagnostic categories as well as to account for the heterogeneous nature of the different task paradigms included in the present analysis (for a similar approach see (37).

## Results

Literature search, screening, and evaluation according to our criteria resulted in a total of 144 articles. In line with our recent meta-analysis (46) using the same SUD dataset to determine common and substance-specific neurofunctional alterations across SUDs and revealing convergent alterations in fronto-striatal regions across different SUDs, data from studies of different SUDs were included (Alcohol, 19.31%; Cocaine, 15.86%; Cannabis, 15.17%; and Nicotine, 12.41%) and pooled for the present analysis. IGD and OCD each contributed to 13.1% and 24.13% of the included studies, respectively. The RDoC domain classification yielded four categories: Positive Valence Systems (PVS), Cognitive Systems (CS), Negative Valence Systems (NVS), and Social Processes (SP), the distribution of the various domains across each disorder shown in **Table S8**. There was no publication bias in the reported sample size related to the reported foci (p=0.45, intercept =0.29, **Fig S2**). There were no significant differences among the four RDoC domains for the three disorders as examined by chi-square tests (PVS: *χ*^2^ = 24, p = 0.24, CS: *χ*^2^ = 20, p = 0.24 NVS: *χ*^2^ = 12, p = 0.28, and SP *χ*^2^ = 18, p = 0.26). In total, data from n = 3494 patients (mean age = 31.76, SD=11.54) and n = 3403 controls (mean age = 29.80, SD = 11.10) were included (details see also the PRISMA flow diagram provided in **Fig 1**).

### Disorder-specific meta-analyses and contributions of RDoC domains

The first meta-analyses examined data from the three diagnostic categories separately to determine clusters most likely determined for each diagnostic group relative to the healthy reference groups. Moreover, the percentage distribution of the classified RDoC domains was computed for each cluster in each diagnostic category. Resembling the results from our previous SUD-focused meta-analysis (46) the main effect across all SUDs primarily revealed clusters located in (dorsal) striatal regions as well as frontal and insular regions (**Fig 2A1, A2**). For all clusters contribution of cognitive systems (RDoC construct: attention, cognitive control, working memory, and memory encoding) were the highest, followed by contributions of the positive valence systems (RDoC construct: approach/motivation and reward attainment). For OCD robust alterations were observed in prefrontal and insular regions, although a similar pattern of RDoC contribution as in SUD was observed (**Fig 2 B1, B2**). For IGD, robust alterations were determined in a widespread network encompassing cortical midline and temporal regions, including superior and middle frontal regions as well as middle, inferior and superior temporal regions, the posterior cingulate and lingual gyrus. With respect to the RDoC domain contributions, cognitive systems, and positive valence systems contributed equally to the alterations in IGD with some contributions from negative valance system (RDoC contrast: acute threat, i.e. fear conditioning or symptom provocation, frustrated non-reward including monetary delay paradigms with punishment) (**Fig 2 C1, C2**). Note that across all disorder categories most original studies employed paradigms assessing cognitive domains and positive valence systems, yet the distribution and contribution of the domains to the identified clusters did not significantly differ Table S8 shows the values associated with the RDoCs.

**Figure 2.**
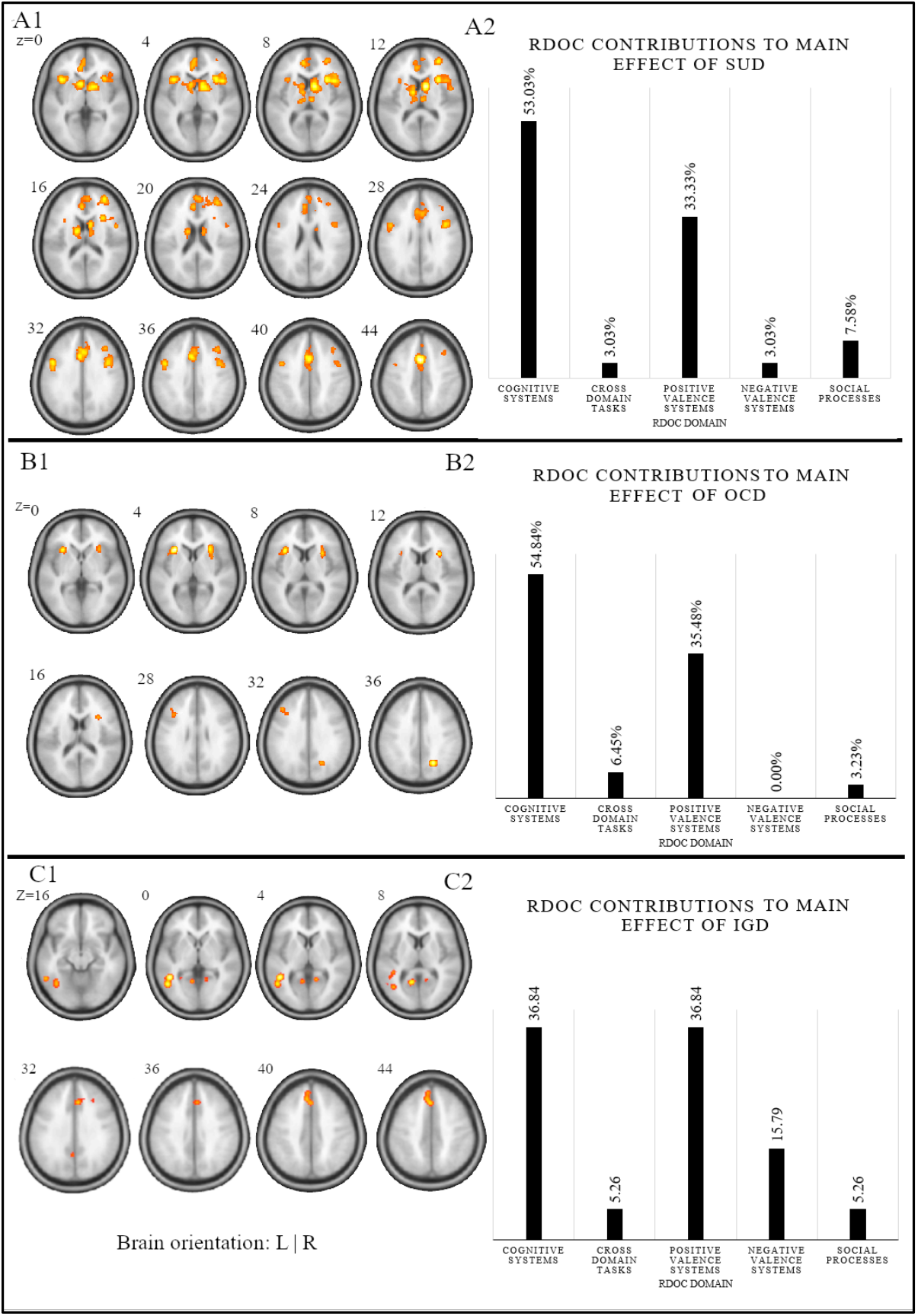
Disorder-specific meta-analysis. Separate meta-analyses on the case-control studies in the three disorders revealed A1) the altered regions activated across all SUD studies, B) the RDoC classification contributing to the altered regions with percentage of the contribution in the bars, B1) the altered regions activated across all OCD studies. B2) the RDoC classification contributing to the altered regions with percentage of the contribution in the bars, C1) the altered regions activated across all IGD studies. C2) the RDoC classification contributing to the altered regions with percentage of the contribution in the bars. Results thresholded at cluster-level FWE, p< 0.05 and initial cluster forming threshold p<0.001. The actual values of the percentages are shown in Table S8

### Meta-analytic determination of differences between the diagnostic categories

A contrast approach was employed to determine significant meta-analytic differences between the three diagnostic categories by comparing the meta-analytic maps of two disorder categories. The corresponding spatial maps represent areas exhibiting stronger alterations in one diagnostic category as compared to the other diagnostic category. Comparing SUD and IGD meta-analytic maps revealed greater alterations in SUD in dorsal striatal and prefrontal as well as anterior cingulate and insular regions, whereas IGD showed greater alterations in temporal regions as well as the middle frontal gyrus in direct comparison with SUD (**Fig 3 A1, Table S2**). The identified regions exhibited mainly contributions of cognitive and positive valence RDoC systems for both, SUD and IGD, respectively (**Fig 3 A2, A3**). The direct comparison of SUD with OCD (**Fig 3 B1, Table S3**) revealed relative greater alterations in SUD in dorsal striatal and prefrontal areas, as well as anterior cingulate and thalamic regions whereas OCD showed greater neurofunctional alterations in posterior frontal and parietal regions. Positive valence and cognitive systems contributed to the alteration in SUD and OCD, respectively (**Fig 3 B2, B3**). A meta-analytic comparison of IGD and OCD revealed stronger alterations in the bilateral insula, posterior parietal, and middles frontal regions in OCD relative to IGD, whereas IGD exhibited stronger alterations in inferior and middle temporal, cingulate and hippocampal regions (**Fig 3 C1**). Alterations in OCD were mainly contributed by cognitive and to a lesser extent positive valence RDoC systems, whereas alterations in IGD were mainly contributed by negative and positive valence systems (e.g. particularly from reward attainment or motivation and delay incentive leading to punishment representing positive valence and negative valence systems respectively) (**Fig 3 C2, C3, Table S4)**.

**Figure 3.**
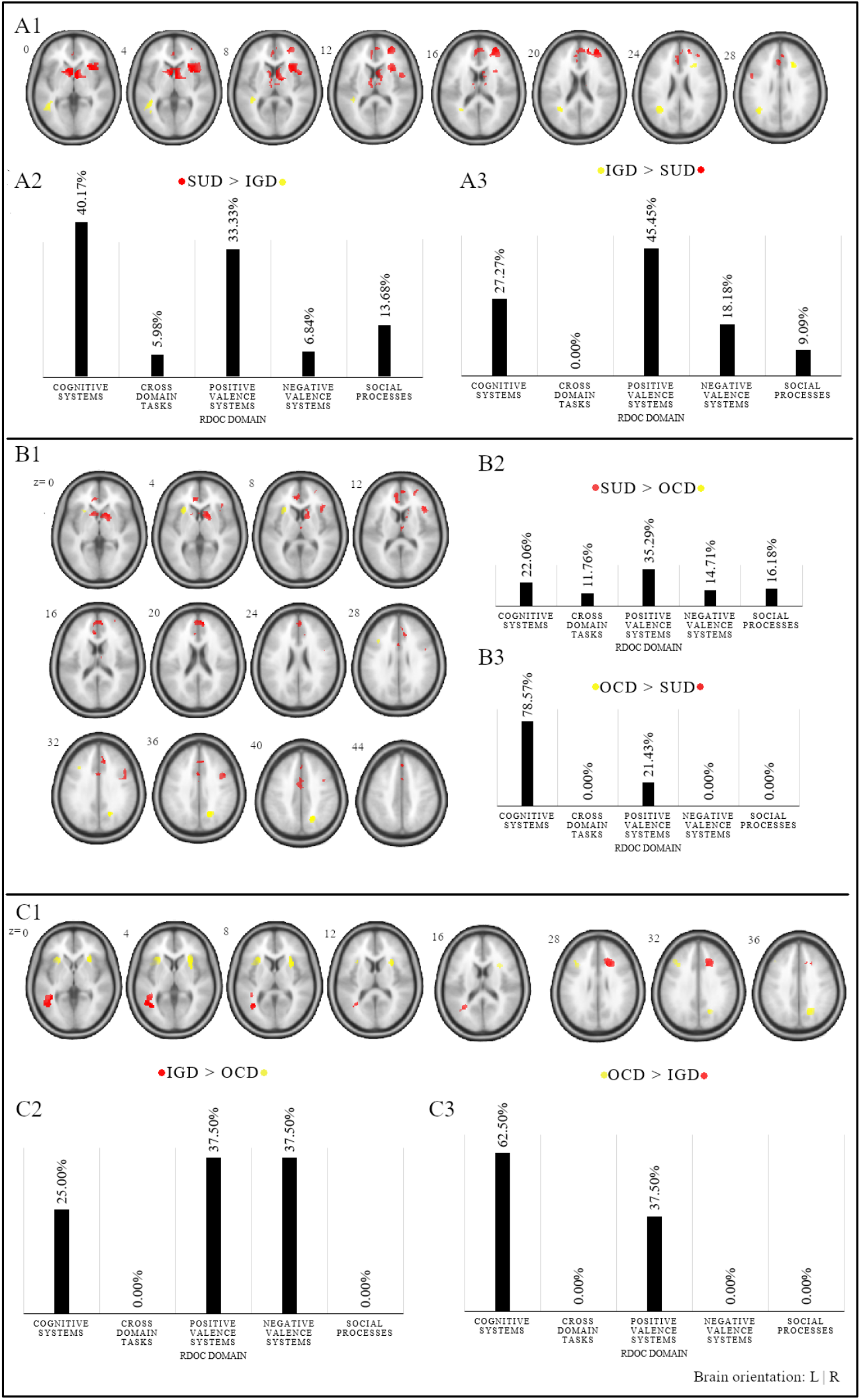
Meta-analytic comparison of alterations in each diagnostic group A1) contrast reflecting relative differences between SUD and IGD A2) the RDoC percentage contribution to pronounced alterations in SUD relative to IGD (clusters N=7), and A3) the RDoC percentage contribution to pronounced alterations in IGD relative to SUD (clusters N=4). Color codes: Red = stronger alterations in SUD, Yellow = stronger alterations in IGD. B1) contrast reflecting relative differences between SUD and OCD B2) the RDoC percentage contribution to pronounced alterations in SUD (clusters N=10), and B3) the RDoC domain percentage contribution to stronger alterations in OCD relative to SUD (clusters N=3). Color codes: Red = stronger alterations in SUD, Yellow = stronger alterations in OCD. C1) contrast reflecting relative differences between IGD and SUD C2) the RDoC percentage contribution to pronounced alterations in (clusters N=2), and C3) the RDoC domain percentage contribution to stronger alterations in IGD relative to OCD (clusters N=3). Color codes: Red = stronger alterations in IGD, Yellow = stronger alterations in OCD. Results thresholded at cluster-level FWE, p< 0.05 and initial cluster forming threshold p<0.001

### Meta-analytic conjunction analyses: shared neurofunctional alterations

The conjunction analysis between SUD and IGD revealed shared alterations in prefrontal regions, specifically in the medial frontal gyrus spreading into the adjacent anterior cingulate and the inferior frontal gyrus (**Fig 4 A1, Table S5**) which were predominately contributed by the cognitive systems RDoC domain (**Fig 4 A2**). The conjunction between SUD and OCD revealed shared alterations primarily in the bilateral anterior insular cortex and the precentral gyrus which were mainly contributed by the RDoC cognitive systems domain and to a lesser extent the positive valence domain (**Fig 4 B1, B2)**. The detailed peaks are presented in **Table S6**. The conjunction analysis between IGD and OCD did not reveal regions exhibiting consistent meta-analytic alterations across the two diagnostic categories.

**Figure 4.**
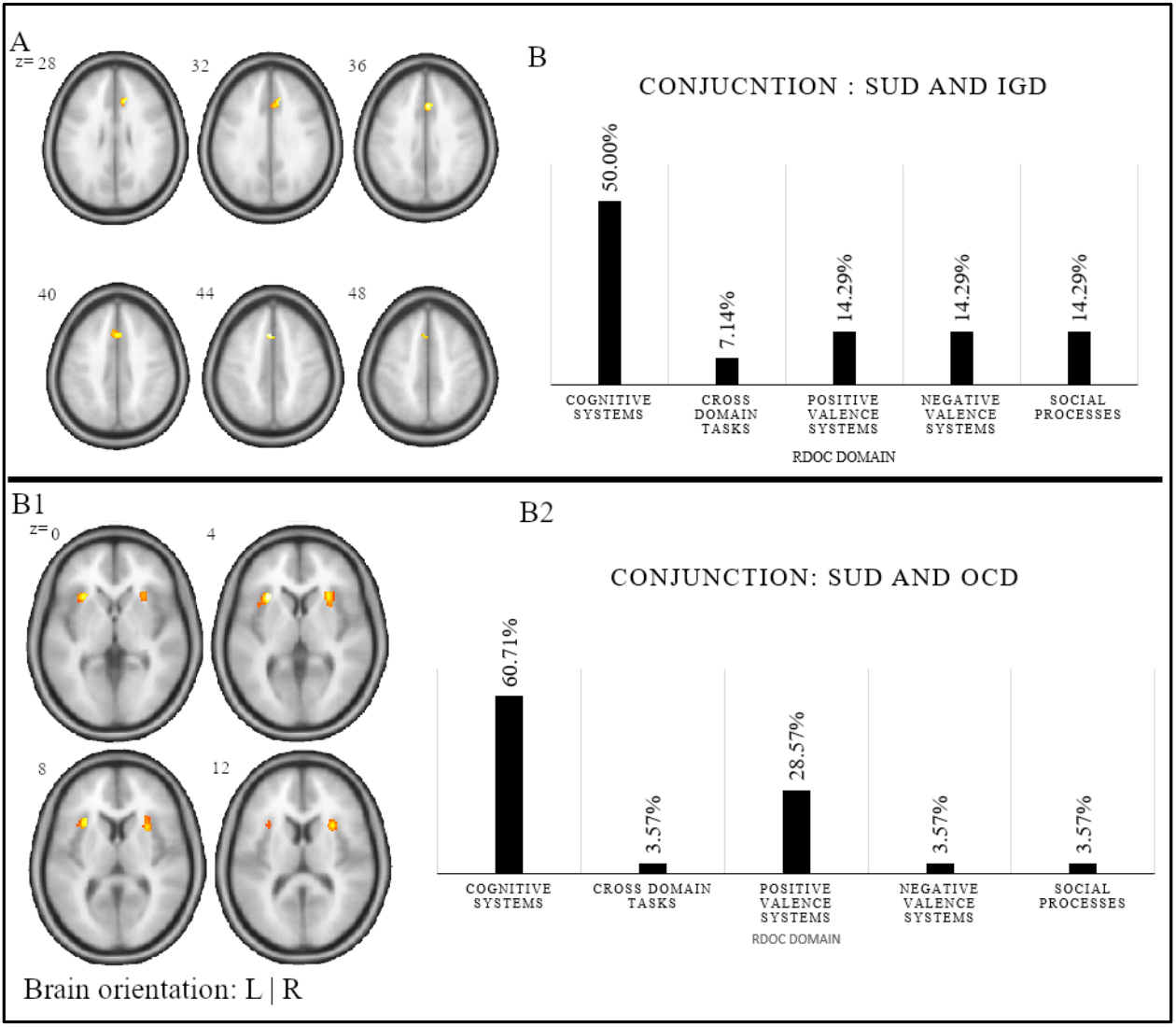
Meta-analytic conjunction analysis A1) Shared alterations between SUD and IGD, A2) percentage distribution of the RDoC contribution to the clusters (N=2). Results thresholded at cluster-level FWE, p< 0.05. B1) Shared alterations between SUD and OCD, B2) the percentage distribution of RDoC contribution to the clusters (N=3). Results thresholded at cluster-level FWE, p< 0.05 and initial cluster forming threshold p<0.001

### Exploratory analysis: contribution of separate SUDs and RDoC domains

Separate meta-conjunction analyses between each SUD (Cocaine, Cannabis, Alcohol and Nicotine) and OCD/IGD respectively revealed highly consistent shared neurofunctional alterations across the SUDs. Specifically, OCD exhibited shared regional-specific neurofunctional alterations of the anterior insular cortex with all SUDs, while IGD exhibited shared neurofunctional alterations of the dorsal anterior cingulate cortex (dACC) with all SUDs **Fig S1**), reflecting a high consistency across SUDs and emphasizing general rather than substance-specific shared alterations. Finally, to examine whether the RDoC domain targeted by the case-control studies may have affected the determination of the differential and shared alterations we examined the heterogeneity of the functional paradigms by means of extracting the voxel-wise probabilities for the clusters from the conjunction analysis and subjecting these to Kruskal-Wallis tests. No significant influence of the RDoC domain categorized paradigms on the conjunction (common) or disorder-specific neurofunctional alterations was observed following adjustment for multiple comparisons (**Table S8**). Moreover, we tested the RDoC distribution for the separate SUDs in comparison with IGD and OCD respectively. Again, the Chi-square test for statistical independence for the individual SUDs argued against the influence of the different paradigm classes on the observed results (*χ*^2^ = 14.5, p = 0.26).

## Discussion

Based on symptomatic overlap and emerging transdiagnostic models for OCD, SUD, and IGD, we aimed at systematically determining shared and disorder-specific neurofunctional alterations between these psychiatric disorders. To this end, we employed a series of coordinate-based meta-analyses that capitalized on a large number of previous case-control fMRI studies in these diagnostic entities. Separately examining neurofunctional alterations within each diagnostic category revealed robust alterations in insular-prefrontal circuits in SUD and OCD, with SUD exhibiting additional alterations in (dorsal) striatal regions, while IGD was characterized by widespread alterations in temporal and posterior cingulate regions as well as middle frontal regions. Across the three diagnostic entities, the identified neurofunctional alterations were predominately contributed by task-paradigms tapping into the cognitive and positive valence systems RDoC domains. In line with the descriptive findings from the separate meta-analyses, direct comparisons between the diagnostic categories revealed that SUD individuals exhibited pronounced dorsal striatal and prefrontal alterations as compared to both, OCD and IGD, whereas OCD subjects exhibited pronounced neurofunctional alterations in the anterior insular cortex (and left dlPFC) and IGD presented pronounced alterations in widespread temporal regions as compared to both other diagnostic categories. Finally, examining the common alterations via meta-analytic conjunction analyses revealed shared alterations between SUD and OCD in the bilateral anterior insular cortex and the precentral gyrus. Subsequent SUD-specific conjunction analyses further demonstrated robust shared neurofunctional alterations between OCD with each SUD category suggesting general rather than substance-specific shared alterations. SUD and IGD shared neurofunctional alterations in right inferior frontal and bilateral medial frontal regions, with substance-specific meta-analysis suggesting convergent alterations between IGD and each SUD in the dorsal anterior cingulate. In contrast to our expectations, no robust shared alterations between OCD and IGD were observed which indicates that IGD, despite having compulsivity as a shared behavioral commonality with OCD, is closer to SUD from functional neuroimaging perspectives. Across analyses and diagnostic categories, the alterations were predominantly contributed by the RDoC cognitive systems domain and to a lesser extent the positive valence domain. One exception was observed with respect to the pronounced temporal alterations in IGD which were partly contributed to by the RDoC negative valence system and by a possible indirect role of the temporal lobe in impulse dysregulations.

The results from the initial meta-analyses that aimed at mapping robust neurofunctional alterations within each disorder category generally align with previous disorder-specific meta-analyses. In line with our SUD-focused previous meta-analysis (46) individuals with chronic substance abuse exhibited robust alterations in striatal, primary dorsal striatal (47), and frontal regions. These findings resonate with several previous meta-analytic findings in substance users demonstrating altered striatal processing of drug-associated and non-drug associated rewards (20,21,46,48) and altered frontal processing during executive and regulatory functions (49,50). Specifically, neurofunctional alterations in the dorsal striatum have been associated with the development of compulsive drug use in humans (51,52) and animal models demonstrated that drug-induced neuroplastic alterations in cortico-striatal circuits critically mediate the formation of habitual and ultimately compulsive patterns of drug use (15,16). In line with previous meta-analyses covering case-control fMRI studies in OCD, we observed robust alterations in frontal and insular networks, primarily during cognitive and positive valence paradigms, in OCD. However, in contrast to these previous meta-analyses, no robust alterations in striatal regions were observed in the present meta-analysis (19,53,54). Partly resembling previous meta-analyses in IGD we observed robust neurofunctional alterations in frontal and temporal regions in IGD subjects, yet no robust alterations in striatal regions (34,55,56) however, see also (57).

The major aim of the present study was to systematically determine shared and disorder-specific neurofunctional alterations between these three diagnostic and symptomatic categories. Directly comparing the meta-analytic maps revealed that SUD was characterized by pronounced alterations in dorsal striatal-prefrontal circuits compared to both, OCD and IGD. These findings emphasize the importance of drug-induced neurofunctional alterations in these circuits which may promote the transition to addictive and ultimately compulsive use. OCD presented stronger alterations in the anterior insular cortex compared to both other diagnostic categories and neurofunctional alterations in the anterior insular cortex were shared between SUD and OCD. The anterior insular cortex has been implicated in a variety of functions ranging from basal interoceptive functions, affective empathy and salience processing to higher-order cognitive functions such as risky decision making (58–61). Lesion studies have demonstrated a critical role of the anterior insula cortex in interoceptive accuracy and drug craving (62), suggesting that the insula integrates bodily sensations with emotion and motivation and thus mediates the conscious awareness of urges (63). In contrast to cortico-striatal circuits, the insula has only recently been conceptualized as a core region of the circuitry that promotes the development of compulsive behavior, with initial results from animal lesion models suggesting a causal role of the anterior insula in the formation of compulsive behavior (64). In contrast to both other diagnostic categories IGD was characterized by widespread neurofunctional alterations in the temporal cortex and to a lesser extend frontal regions. Temporal lobe regions have not been robustly identified in previous meta-analyses of either OCD or SUD (19,21,46,53,54,65,66), while some previous meta-analysis on IGD reported temporal as well as frontal alterations, specifically during higher-level cognitive processes such as executive control (34,55). The findings of IGD-specific alterations in temporal lobe regions are in line with initial studies comparing IGD and SUD samples reporting opposing intrinsic communication alterations between prefrontal regulatory control nodes and temporal regions in IGD and SUD (67) as well as a recent report suggesting opposite associations between impulsivity and temporal lobe thickness in IGD and SUD (67,68), thus further emphasizing the possible regulatory role of temporal lobe in impulse dyscontrol by its interaction with the prefrontal cortex.

In line with several previous cross-sectional meta-analytic and prospective studies reporting functional and structural alterations in IGD (34,52,55,56) the present meta-analysis found altered prefrontal activation in IGD, of which alterations in inferior frontal and medial frontal regions were shared with SUD. The integrity of these prefrontal cortex regions critically mediates executive and inhibitory control functions and the shared alterations may reflect marked deficits in these domains as previously reported in individuals with both disorders (69,70).

In contrast to our hypotheses, IGD did not demonstrate robust shared neurofunctional alterations with OCD. Together with the overlapping prefrontal alterations between IGD and SUD as well as the pronounced temporal alterations in IGD relative to both disorders the pattern of results may indicate that prefrontal deficits potentially related to common inhibitory and executive deficits represent shared neurofunctional alterations across substance and behavioral addictions, while neural alterations in the anterior insular cortex potentially related to compulsivity are shared between substance addiction and OCD yet do not represent a characteristic of IGD. These findings may indicate that the compulsive symptoms observed on the behavioral level in IGD are either less dominant in this disorder or mediated by different neuropathological pathways.

Although the present meta-analysis revealed important insight in common and distinct neurofunctional alterations across the disorders, the influence of potentially confounding factors could only be analyzed to a certain extent due to the nature of the original studies. For example, studies within each disorder group presented different indices of substance use such that some studies recorded the duration of dependency for substances while others reported the amount of substances or medication used or no corresponding indices. Moreover, the original studies included in this meta-analysis employed cross-sectional case-control designs, which limits conclusions of whether the observed neurofunctional alterations represent pre-disposing risk factors or represent a consequence of the underlying disorder. Finally, to increase statistical power to determine shared neurofunctional alterations the present analysis only considered relative alterations from previous case-control studies without accounting for reduced or increased activation in the patients relative to the healthy reference group. The corresponding information could reveal important information for pathology models and should be included in subsequent meta-analysis that can capitalize on a larger number of studies.

## Conclusion

To our knowledge this is the first study that takes advantage of the currently available fMRI case-control studies to determine common and separable neurofunctional alterations in SUD, OCD, and IGD, thus taking an important step forward in this rather unexplored territory. The alterations within each diagnostic category emphasize that neurofunctional alterations in frontal regions characterize the disorders, with OCD and SUD presenting pronounced alterations in the insula, and additional dorsal striatal alterations in SUD. IGD and SUD exhibited shared neurofunctional alterations in prefrontal regions engaged in inhibitory and executive control, while IGD and SUD did not exhibit common neurofunctional alterations. The identified neurofunctional alterations across the three diagnostic entities were predominately contributed by task-paradigms tapping into the cognitive and positive valence systems RDoC domains and may suggest shared cognitive and reward-related processing behavioral alterations underlying the three disorders. If replicated in future studies, this may pave the path towards neuroimaging informed targeted interventions such as non-invasive brain stimulation/neuromodulation interventions for these often chronic and difficult to treat conditions.

## Data Availability

The data is available from the corresponding authors upon reasonable request.

## Authors contribution

The contributions of the authors involved in this study are as follows: research conceptualization and methods, drafting/writing manuscript, and statistical/formal analysis. Klugah-Brown; conceptualization analysis, drafting of the manuscript, Zhou; acquisition and inspection of data, Zweerings; interpretation final approval of analysis and manuscript, Mathiak; validation and revision of interpretation and analysis, Biswal; revision and approval of manuscript and Becker; conceptualization, interpretation and final approval of the manuscript.

## Data availability statement

The data is available from the corresponding authors upon reasonable request

